# Development and Psychometric Evaluation of the AASPIRE Social Support Scale for Autistic Adults

**DOI:** 10.64898/2026.10.29.26366606

**Authors:** Wei Song, Todd C. Edwards, Mary Baker-Ericzén, Willi Horner-Johnson, Rachel Ludwig-Kripke, Steven K. Kapp, Katherine E. McDonald, Ian Moura, Liu-Qin Yang, Julie Lounds Taylor, Joseph W. Vera, Christina Nicolaidis

**Author notes:** **Funder Information** National Institute of Mental Health, https://ror.org/04xeg9z08, R01MH121407.

## Abstract

**Background:** Social support is associated with health, well-being, and quality of life among autistic adults, yet existing social support measures have important limitations. Many were not developed with autistic people, may be difficult for autistic adults to use, and have limited psychometric evidence in autistic populations. We evaluated the psychometric properties of the AASPIRE Social Support Scale, an autism-informed measure adapted from Patient-Reported Outcomes Measurement Information System (PROMIS) social support scales through a community-based participatory research process.

**Methods:** The AASPIRE Social Support Scale assesses five domains: Companionship, Emotional Support, Informational Support, Practical Support, and Social Isolation. Baseline data were analyzed from 859 autistic adults recruited through healthcare systems, disability service systems, and the broader U.S. autism community. Participants completed the measure independently (n=600), with support (n=189), or through caregiver report (n=70). We evaluated content validity, structural validity using confirmatory factor analysis, internal consistency, and measurement invariance across reporting approaches. We also explored differences in social support scores by reporting approach and participant characteristics.

**Results:** The hypothesized five-factor structure demonstrated good fit, with strong item loadings supporting the five distinct but related social support domains. Internal consistency was high for the total scale and all subscales (Cronbach’s α=.87–.95). Measurement invariance was supported across independent direct-report, supported direct-report, and caregiver report, indicating that the scale measured the underlying social support domains similarly across reporting approaches. Social support scores differed across reporting groups: participants reporting independently generally reported less support and greater isolation than participants completing the measure with support or through caregiver report. Exploratory analyses also identified differences by age and gender, but not race.

**Conclusions:** The AASPIRE Social Support Scale is an accessible, reliable, and valid multidimensional measure of social support for autistic adults. Its comparable measurement properties across independent direct-report, supported direct-report, and caregiver report support its use in studies seeking to include autistic adults with diverse communication and support needs. The scale may be useful for evaluating social support and the effectiveness of services and interventions across heterogeneous autistic adult populations.

## Introduction

Social support is a multidimensional construct that encompasses both the functions provided by social relationships and individuals’ perceptions of the availability and adequacy of those functions(Sarason et al., 1987; Taylor, 2011). It is a crucial social determinant of health for both the general population and disabled people. It plays a vital role in helping individuals meet their physical and emotional needs and mitigates the adverse effects of stressful life events (e.g.,(Chen et al., 2023; Fisher et al., 2022; Hendryx et al., 2009; Scott & Havercamp, 2014). Among autistic adults, multiple studies have linked more and higher quality social support to reduced suicidality(Hedley et al., 2017; Mournet et al., 2023), less loneliness(Ee et al., 2019; Hedley et al., 2018), and better quality of life (QoL)(Bishop-Fitzpatrick et al., 2017; Charlton et al., 2023; Kamio et al., 2012; Khanna et al., 2014; Mason et al., 2018; Renty & Roeyers, 2006). Further, feeling unsatisfied with social support contributed to perceived barriers to health care among autistic adults(Arnold et al., 2024).

Researchers have differentiated types of *functional* support(Cohen & Wills, 1985; Wills, 1991), such as informational support (e.g., providing advice, feedback, and information to help individuals understand situations and make decisions), instrumental or practical support (e.g., providing tangible assistance with tasks or everyday needs), and emotional support (e.g., showing care, understanding, and compassion). Additionally, scholars have identified social companionship, which involves spending quality time with others and engaging in recreational activities, as a distinct and meaningful form of support (Cohen & Wills, 1985). Importantly, perceived social support is conceptually distinct from support actually received and demonstrated a more consistent association with health outcomes than amount or frequency of social contacts (Sarason et al., 1987; Taylor, 2011). More recent conceptualizations of social support have argued that subjective social isolation, which reflects individuals’ perceptions of whether the quantity and quality of their social relationships adequately meet their needs, is a closely related construct to social support (Wang et al., 2018). Together, these perspectives support a multidimensional conceptualization of social support that differentiates the perceived availability of different functional forms of support, including companionship, emotional, informational, and practical support, while also incorporating perceived social isolation as a complementary appraisal-based domain.

Existing measures used with autistic adults reflect different conceptualizations of social support. The Multidimensional Scale of Perceived Social Support (Zimet et al., 1988) organizes perceived support according to the source of support (e.g., family, friends, and significant others), while the Social Support Questionnaire (Monteiro, 2011; Sarason et al., 1983) focuses on the availability of supportive others and satisfaction with available support. Although these measures assess important aspects of perceived social support, neither directly differentiates the functions that supportive relationships provide. The Interpersonal Support Evaluation List-12 Item Version (ISEL-12; (Cohen & Hoberman, 1983; Cohen et al., 1985; Merz et al., 2014) more closely aligns with a functional conceptualization by assessing the perceived availability of different functions of appraisal, belonging, and tangible support. The ISEL-12 has been used in multiple studies of autistic people (e.g., Hedley et al., 2017). Autism-specific measures have further broadened how social support is operationalized. For example, Social Network and Perceived Social Support (SNaPSS; (Lei et al., 2019)integrates information about social network structure with perceived support across academic, daily living, and socialization domains. Together, these approaches illustrate that social support can be operationalized in multiple ways, with different measures emphasizing the functions, sources, availability, or structural context of supportive relationships.

Despite the availability of these measures, important questions remain regarding how social support should be measured as an outcome for autistic adults. Most existing instruments were developed without autistic people, and measures developed for the general population may embed assumptions about supportive relationships that do not fully reflect autistic adults’ experiences. Moreover, relatively few social support measures have undergone comprehensive psychometric evaluation in autistic adult populations, limiting confidence in their validity and interpretation in this population. These concerns are particularly important for perceived social support because it reflects an individuals’ subjective appraisal of whether support is available and adequate, making direct self-report important whenever possible. At the same time, autistic adults have described difficulties with survey instruments using ambiguous or complex language, imprecise response options, or questions whose meaning varies across contexts, causing them significant confusion, frustration, and anxiety (Nicolaidis et al., 2019; Nicolaidis et al., 2015; Nicolaidis et al., 2020). Relying exclusively on independent self-report may therefore limit participation among autistic adults who require assistance completing surveys or who cannot provide direct reports even with appropriate accommodations. Measures intended for broad use among autistic adults should therefore prioritize accessible direct reporting, including supported self-report when needed, while also providing carefully developed proxy-report options when direct reporting is not possible.

The multidimensional nature of social support, limited involvement of autistic people in the development of measures, gaps in psychometric properties, and barriers to accessible and inclusive reports provided the rationale for using a community-based participatory research (CBPR) approach to identify and operationalize social support within the Academic Autism Spectrum Partnership in Research and Education (AASPIRE) Outcomes Projects. AASPIRE used a CBPR-nested Delphi process in which autistic community members, researchers, service providers, and other partners identified social support as a high-priority outcome and evaluated candidate approaches to its measurement (Nicolaidis et al., 2025). Through this process, the partnership prioritized a multidimensional approach that assessed the perceived availability of distinct forms of functional support, including companionship, emotional support, informational support, and practical support, while also capturing subjective social isolation as a related social outcome. The Patient-Reported Outcomes Measurement Information System (PROMIS) social health measures provided the closest existing framework for assessing these prioritized domains, with separate item banks assessing Companionship, Emotional Support, Informational Support, Instrumental Support, and Social Isolation (Bevans et al., 2014). The PROMIS measures therefore served as the foundation for the AASPIRE Social Support Scale and were subsequently adapted through the AASPIRE CBPR process to improve their relevance and accessibility for autistic adults.

### The Present Study

The present study provides an in-depth psychometric evaluation of the AASPIRE Social Support Scale, developed as part of the larger AASPIRE Outcomes Project. The resulting measure consists of five subscales—Companionship, Emotional Support, Informational Support, Practical Support, and Social Isolation—and includes direct-report and caregiver-report versions to support participation across a range of communication and support needs. Initial psychometric evaluation conducted as part of the broader AASPIRE Measurement Toolkit study demonstrated high accessibility and completion rates, strong two-week test–retest reliability (ICC = .91 for the total scale; ICCs = .82–.91 for the subscales), and responsiveness to change, with changes in subscale scores corresponding to participants’ perceived changes in the respective social support domains (Nicolaidis et al., 2026). Building on this initial evidence, the present study evaluates the measurement properties of the AASPIRE Social Support Scale in greater depth. Specifically, we examine content validity informed by the CBPR adaptation process and cognitive interviewing, construct validity using confirmatory factor analysis, measurement invariance across report types (i.e., direct report, direct report with support, and caregiver report), and internal consistency reliability for each subscale and total scale. Following the evaluation of measurement invariance, we also examine differences in social support scores across reporting approaches and conduct exploratory comparisons by age, gender, and race/ethnicity. Together, these analyses provide a more comprehensive evaluation of the scale’s measurement properties and its applicability across reporting approaches.

## Methods

### Development of the AASPIRE Social Support Scale

As part of the CBPR-nested Delphi process described above, team members and outside experts reviewed three candidate instruments to operationalize social support: (1) the Medical Outcomes Study Social Support Survey (MOS-SS), which had been previously adapted for use with people with developmental disabilities(Nicolaidis et al., 2015); 2) the UCLA Loneliness Scale, a commonly used measure to assess the subjective experience of social disconnection(Grace et al., 2022); and 3) the PROMIS item bank for companionship, emotional, instrumental, and informational support and isolation. The Delphi panel evaluated their coverage of the intended constructs, relevance to autistic adults, and anticipated accessibility. Of the candidate measures, Delphi panelists felt that the PROMIS short forms best captured the intended construct and would require the least adaptation to make them accessible to autistic adults.

In the next phase of the project, we worked in a community-academic workgroup to adapt the selected measures for autistic adults. We reviewed individual items to improve their clarity, language accessibility, and conceptual fit and presented proposed adaptations to the full community-academic team for iterative feedback. Adaptations included adding prefaces to each subscale; adding visuals to all response options (cylinders filled to different proportions); modifying text to clarify terms and concepts; and substituting some items with other items from the PROMIS item banks or with new items we created to better capture autism-related dimensions of social support. For the Practical Support subscale, we added hotlinks with more detailed instructions for how to answer if participants do not currently need support for the activities described in the items and added a fifth item asking about whether someone would notice and step in if they needed help.

The adapted measure retained five domains represented in the selected PROMIS measures: Companionship, Emotional Support, Informational Support, Practical Support, and Social Isolation. To further establish content validity and accessibility, the broader toolkit development process included iterative cognitive interviews with 37 autistic adults and caregivers, 8 of whom reviewed the social support scale and identified only minimal need for further refinement.

Information regarding the adaptation process and cognitive interviewing procedures is reported elsewhere(Nicolaidis , 2026). Supplemental Material 1 includes the final versions, as well as a list of specific changes from the original PROMIS scales.

### Study Sample

Participants were drawn from the AASPIRE Outcomes Project longitudinal cohort study. We recruited autistic adults from two healthcare systems, two disability service systems, and the broader U.S. autism community to ensure a heterogeneous sample and support validation across real-world settings. Eligible participants were at least 18 years old, resided in the United States, and identified as autistic. Participants in the healthcare and disability services sub-cohorts also had to have a formal diagnosis of autism in their health or agency records, respectively. Detailed information about the cohort, recruitment procedures, eligibility criteria, and participant characteristics is reported in Nicolaidis et al. (2026).

The present analyses included baseline data from 870 participants. Eleven participants were excluded because they had complete missingness in social support items. The remaining 859 participants were included in the final analysis. The missingness in individual items of the AASPIRE Social Support Scale ranged from 0.1% to 1.0% among the 859 participants.

### Data Collection Procedure

We collected data between April 2023 and December 2024. Participants could complete surveys directly, with or without support from another person. When direct participation was not possible even with support, a caregiver could complete a caregiver-report version on the autistic participant’s behalf. Participants could complete the survey online or with a research assistant via telephone, videoconference, or in-person when feasible. The present study used baseline data, which provided the largest available sample for evaluating the scale’s factor structure, internal consistency, measurement invariance, and differences in social support across reporting approaches and participant characteristics. The AASPIRE Social Support Scale was administered as part of the broader AASPIRE Measurement Toolkit. A detailed description of participant recruitment, informed consent procedures, scammer prevention procedures, cohort design, accessibility accommodations, longitudinal follow-up procedures, and data management methods is available in Nicolaidis et al. (2026).

### Analysis Strategy

We used the COSMIN guidance (Mokkink et al., 2010) to establish validity and reliability of the AASPIRE Social Support Scale. First, to assess structural validity, we used confirmatory factor analysis (CFA) to test the pre-specified five-factor structure corresponding to the five subscales: Companionship, Emotional Support, Informational Support, Practical Support, and Social Isolation. CFA was conducted in Mplus 8.11 (Muthén & Muthén, 2024) using robust maximum likelihood (MLR) estimator. After excluding a small subset with complete missingness on all social support items, we analyzed the remaining sample using full information maximum likelihood (FIML) to account for item-level missing data. The criteria used to judge model fit included the comparative fit index (CFI > .90; Kline, 2005), the root-mean-square error of approximation (RMSEA < .07; Kline, 2005; Steiger, 2007), and the standardized root-mean-square residual (SRMR <.10; Kline, 2005).

After establishing structural validity, we calculated Cronbach’s alpha to evaluate internal consistency for each subscale, overall and stratified by reporter type (self, self with support, or caregiver). We also reported McDonald’s omega as a measure of reliability for multi-point scales (McNeish, 2018). Omega coefficients are recommended in congeneric scales where items have unequal relations to the construct being measured, such as in a factor analysis setting (Raykov, 1997).

We tested measurement invariance to determine if the scale functions equivalently across reporter types. Three nested models were tested and compared, including a configural model where parameters were free to vary, a metric model where factor loadings were constrained to be equal, and a scalar model where factor loadings and intercepts were constrained to be equal across groups. To achieve strong measurement invariance that allows the comparisons of latent mean differences across groups, equal factor loadings and equal item intercepts across groups must be in place (Cheung & Rensvold, 2002; Widaman & Reise, 1997). For the model comparison, the Chi-square difference test and changes in alternative fit indices were used to evaluate factorial invariance. Specifically, a non-significant change in chi-square values and trivial changes (.01 or less) in other fit indices, including CFI, RMSEA, and SRMR, were considered as evidence of invariance (Chen, 2007; Cheung & Rensvold, 2002). Nested model comparisons were evaluated using the Satorra–Bentler scaled χ^2^ difference test for robust estimators.

After establishing measurement invariance across the reporter types, we further compared social support total scale score and subscale scores by reporter types. We also conducted exploratory comparisons by age groups (18-29 vs. others), gender (men, women, and other), and race (BIPOC or not). Given the limited prior literature regarding demographic variation in perceived social support among autistic adults, we did not specify a priori hypotheses regarding these differences; these analyses were intended to characterize score distribution across subgroups and inform hypotheses for future research. One-way ANOVA was conducted to test overall group differences, followed by pairwise comparisons with Bonferroni adjustments for multiple comparisons.

## Results

### Sample Characteristics

About 69.8% (n=600) of them completed the AASPIRE Social Support Scale independently (i.e., direct-reports), 22.0% (n=189) completed with support (i.e., direct-reports with support), and 8.1% (n=70) were completed by a caregiver (i.e., caregiver reports). The average age of the autistic adult sample was 30.9 years (SD=11.59, ranging from 18 to 77), and 28.5% of the sample were Black, Indigenous, or People of Color (BIPOC). About half of the sample found it hard or very hard to afford basic necessities, and almost half of the sample (48.9%) had at least some paid employment. Table 1 presents the participant characteristics.

**Table 1.**
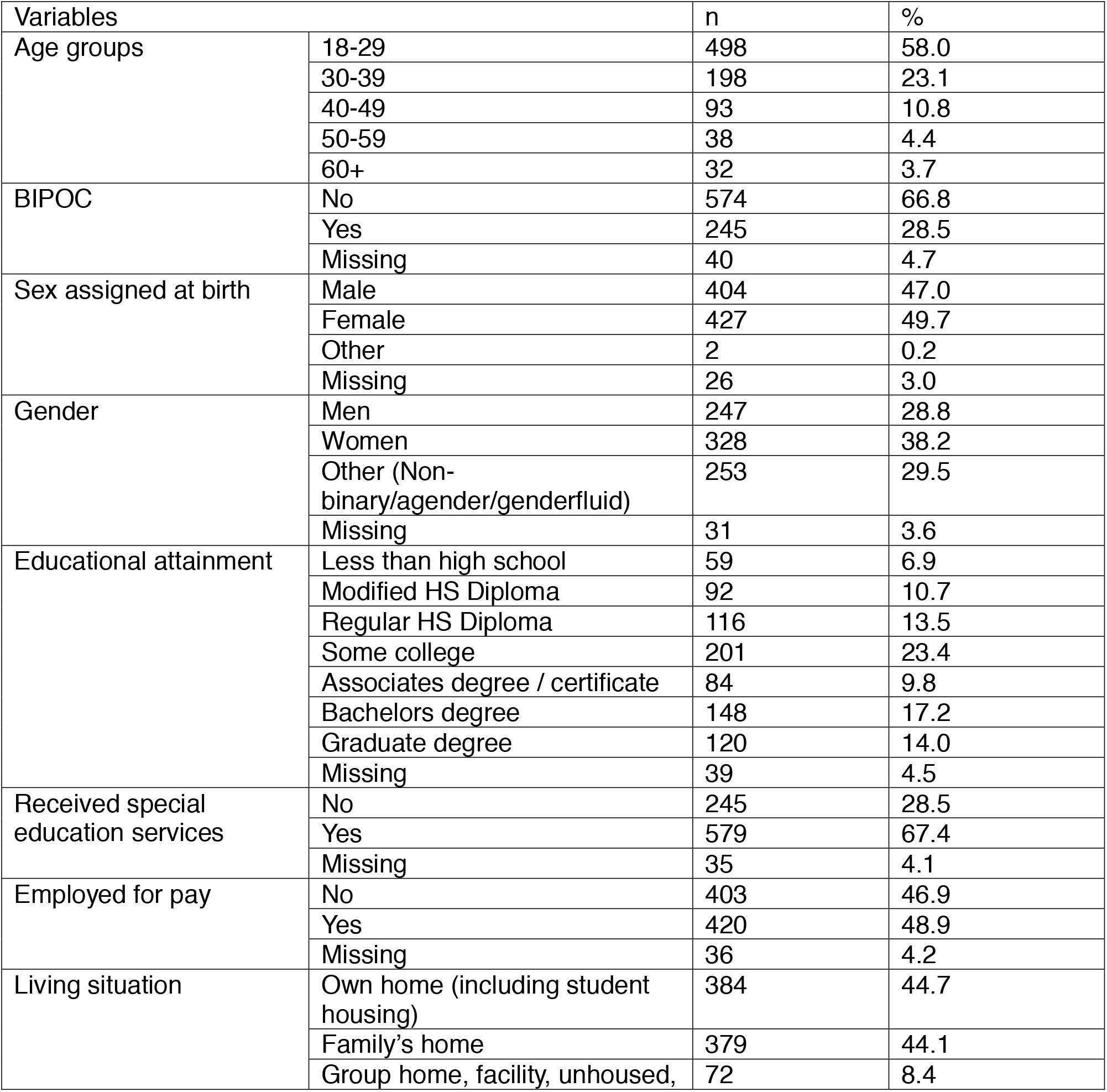
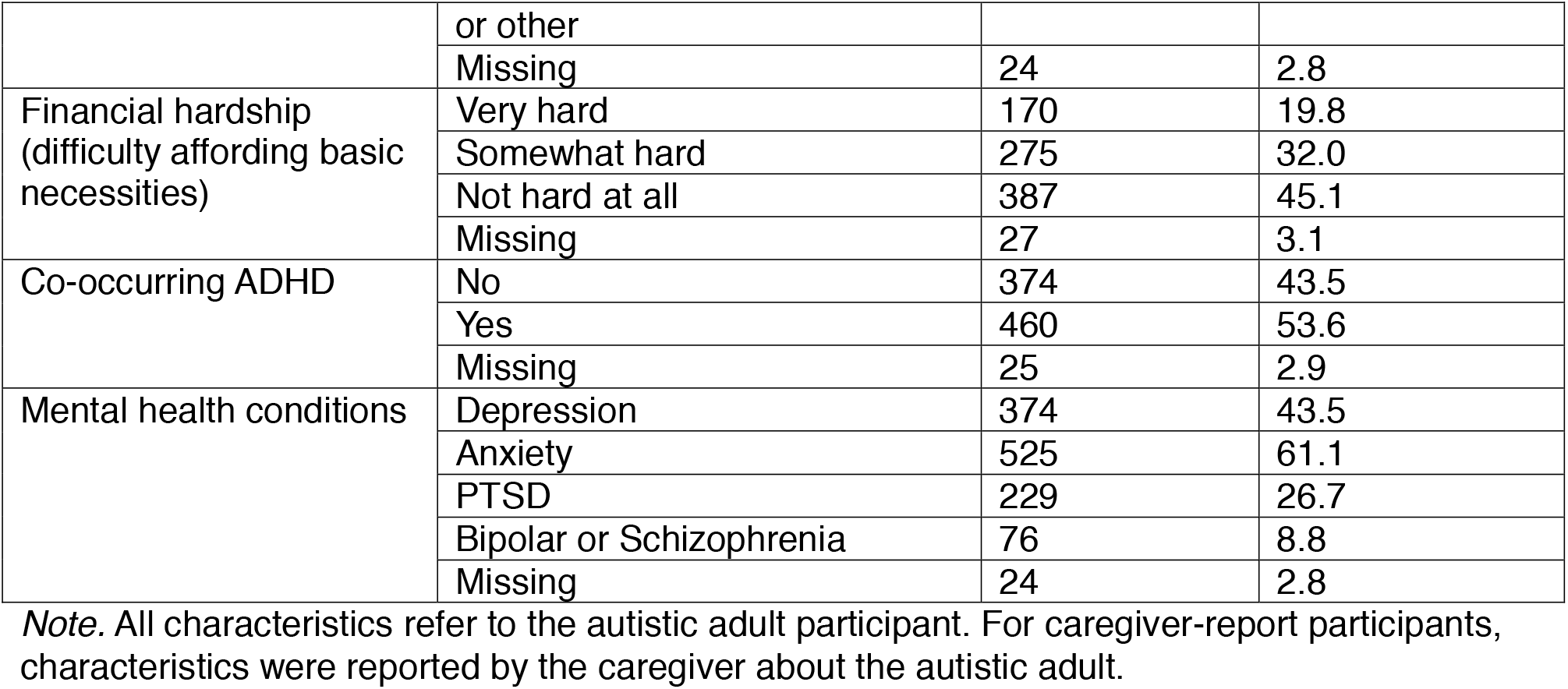
Characteristics of study participants (N=859)

### Structural Validity using CFA

Overall, the hypothesized five-factor structure was well-supported using the full sample, based on very strong item loadings, excellent CFI/TLI, and factor correlations. Modification indices did not suggest substantially meaningful cross-loadings and only indicated trivial correlated residuals. The CFA model indicated that the five-factor model was appropriate for the whole sample: χ^2^(179) = 623.75, CFI = .955, TLI = .947, RMSEA = .054 (90% CI: 0.049, .058), SRMR = .045. Additionally, all standardized factor loadings were strong (ranging from 0.71 to 0.89, all *p* < .001).

Moderate to high correlations were observed between the factors (ranging from 0.35 to 0.77; Table 2). Emotional Support had high correlations with both Companionship (r=0.73) and Informational Support (r=0.77). Practical support had a relatively weak association with social isolation (r=0.35).

**Table 2.** Inter-factor correlations (standardized)

| Factors | 1.<br>Companionship | 2.<br>Emotional | 3.<br>Informational | 4.<br>Practical | 5.<br>Isolation |
| --- | --- | --- | --- | --- | --- |
| 1.<br>Companionship | — |  |  |  |  |
| 2. Emotional | .73 | — |  |  |  |
| 3. Informational | .56 | .77 | — |  |  |
| 4. Practical | .54 | .46 | .49 | — |  |
| 5. Isolation | .51 | .59 | .54 | .35 | — |

*Note.* All values are standardized latent correlations estimated from the five-factor confirmatory factor analysis model. All correlations with $p$ values $<0.001$ .

### Measurement Invariance Across Reporter Types

We evaluated the measurement invariance of the Social Support Scale across three respondent groups (direct-report, direct-report with support, and caregiver proxy report). Results of each model and model comparison are presented in Table 3.

**Table 3.**
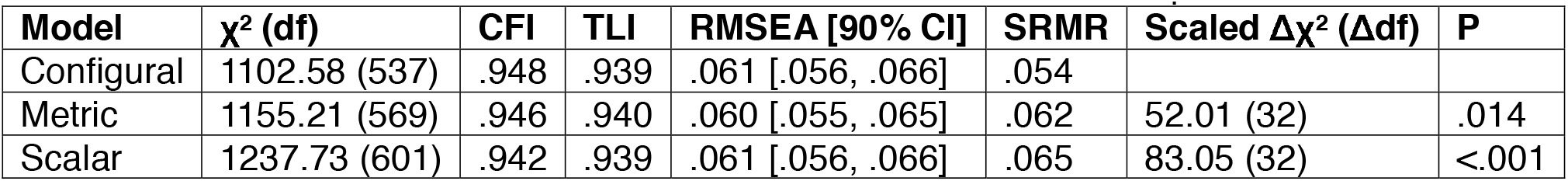
Indices of model fit for measurement invariance and model comparison.

The configural model, in which the factor structure was freely estimated across groups, demonstrated good fit, indicating that the five-factor model was appropriate for all groups, χ^2^(537) = 1102.58, CFI = .948, TLI = .939, RMSEA = .061 [90% CI: .056, .066], SRMR = .054. The **metric model**, which constrained factor loadings to equality across groups, also fit the data well (CFI = .946, TLI = .940, RMSEA = .060), with negligible changes in fit indices (ΔCFI = −.002; ΔTLI= +.001; ΔRMSEA = -.001), supporting metric invariance. The **scalar model**, which further constrained item intercepts to equality, likewise demonstrated minimal deterioration in model fit (CFI = .942, TLI = .939, RMSEA = .061; ΔCFI = −.004; ΔTLI= -.001; ΔRMSEA = +.001), supporting scalar invariance.

Although the Satorra–Bentler scaled χ^2^ difference tests were statistically significant (Metric vs. Configural: Δχ^2^(32) = 52.01, p = .014; Scalar vs. Metric: Δχ^2^(32) = 83.05, p < .001), these results likely reflect the large sample size, as the changes in approximate fit indices (ΔCFI = .001–.004; ΔRMSEA = .001) were well within accepted thresholds (Cheung & Rensvold, 2002; Chen, 2007). Therefore, both metric and scalar invariance were considered to be established, supporting the comparability of the social support scale across direct-report, direct-report with support, and caregiver-report groups.

### Internal Consistency of Social Support Scale and Subscales

Internal consistency estimates for the five subscales and the total social support scale were high across the full sample and consistent across reporting groups (Table 4). Cronbach’s α coefficients ranged from .87 to .95 for the subscales and .93 for the total scale, exceeding the recommended threshold of .70 and indicating high internal reliability. McDonald’s ω coefficients were comparable to α values, further supporting the internal consistency of the scale.

**Table 4.** Internal consistency of the subscales and the full scale.

| Subscale | No. of Items | Cronbach's $\alpha$ | | | | McDonald's $\omega$ | | | |
| --- | --- | --- | --- | --- | --- | --- | --- | --- | --- |
|  |  | Total | Direct reports | With support | Caregiver report | Total | Direct reports | With support | Caregiver report |
| Companionship | 4 | 0.89 | 0.90 | 0.82 | 0.88 | 0.89 | 0.90 | 0.82 | 0.88 |
| Emotional Support | 4 | 0.88 | 0.89 | 0.86 | 0.83 | 0.88 | 0.89 | 0.86 | 0.84 |
| Informational Support | 4 | 0.90 | 0.91 | 0.86 | 0.94 | 0.91 | 0.91 | 0.86 | 0.95 |
| Practical Support | 5 | 0.92 | 0.91 | 0.87 | 0.93 | 0.92 | 0.91 | 0.87 | 0.93 |
| Social Isolation | 4 | 0.87 | 0.86 | 0.86 | 0.94 | 0.88 | 0.86 | 0.87 | 0.94 |
| **Total Scale** | 21 | 0.94 | 0.94 | 0.92 | 0.93 | 0.93 | 0.93 | 0.91 | 0.93 |

### Group Differences in Social Support

As scalar measurement invariance was identified across the reporter types, we were able to compare the social support total score and subscale scores between these groups (Table 5).

**Table 5.** Descriptive statistics on the AASPIRE Social Support Scale and subscale total scores, split by reporter types, age, gender, and race.

|  | **Full Scale M(SD)** | **Companionship M(SD)** | **Emotional Support M(SD)** | **Informational Support M(SD)** | **Practical Support M(SD)** | **Social Isolation M(SD)** |
| --- | --- | --- | --- | --- | --- | --- |
| **Report Types** |  |  |  |  |  |  |
| Direct report without support | 66.54 (15.91) | 12.87 (4.00) | 13.73 (3.84) | 13.65 (3.66) | 15.81 (5.53) | 10.48 (3.65) |
| Direct report with support | 75.54 (14.42) | 14.13 (3.47) | 15.15 (3.68) | 14.95 (3.53) | 19.45 (4.93) | 11.86 (4.07) |
| Caregiver report | 81.3 (13.91) | 15.40 (3.47) | 14.59 (4.06) | 15.09 (4.72) | 23.77 (2.86) | 12.46 (4.36) |
| **Age** |  |  |  |  |  |  |
| 18-29 | 65.67 (17.06) | 12.62 (4.19) | 13.4 (3.91) | 13.32 (3.87) | 15.8 (5.97) | 10.52 (3.72) |
| 30 and above | 72.66 (14.94) | 13.88 (3.64) | 14.62 (3.76) | 14.59 (3.62) | 18.32 (5.39) | 11.25 (3.95) |
| **Gender** |  |  |  |  |  |  |
| Men | 70.11 (16.86) | 13.70 (3.75) | 14.12 (3.99) | 14.16 (3.72) | 17.14 (5.89) | 11 (3.93) |
| Women | 72.34 (16.38) | 13.39 (4.07) | 14.34 (3.94) | 14.34 (3.99) | 18.71 (5.65) | 11.56 (4.09) |
| Other (Non-binary/agender/genderfluid) | 66.07 (14.97) | 12.99 (3.98) | 13.83 (3.70) | 13.58 (3.57) | 15.45 (5.37) | 10.22 (3.34) |
| **Race** |  |  |  |  |  |  |
| BIPOC | 69.19 (16.48) | 13.03 (4.12) | 14.01 (3.94) | 14.08 (3.93) | 17.25 (5.82) | 10.82 (4.12) |
| Other | 70.00<br>(16.20) | 13.48 (3.87) | 14.16<br>(3.85) | 14.04 (3.73) | 17.28<br>(5.80) | 11.05<br>(3.76) |

*Note.* Higher scores indicate greater perceived support; Social Isolation items were reverse-scored such that higher scores indicate less social isolation.

The ANOVA showed significant overall group differences in the total scale score and subscale scores (ps<0.001). Pairwise comparison with Bonferroni adjustments showed that individuals reporting directly without support reported lower levels of companionship, informational support, and practical support; higher isolation; and lower total support than both direct report with support and caregiver reports (p<0.001). Those in the direct report without support category also reported lower emotional support than those in the direct-report with support group (p<0.001), but not caregiver report group (p=0.229). Reports completed by caregivers indicated higher practical support and total social support than the other two groups (p<0.001).

We further conducted an exploratory test using ANOVA to determine if there were any group differences in social support in our sample (Table 5). Young participants (18-29 years old) consistently had lower total scale score and subscale scores than the older participants (all p values<0.01). No BIPOC versus non-BIPOC differences were found in any of these outcomes. There were significant gender differences in practical support, isolation, and total social support (p<0.001). Pairwise comparisons with Bonferroni adjustments showed that the other gender (Non-binary/agender/genderfluid) group had lower total social support and practical support than both men and women (p<0.05). They also reported a higher level of isolation than women (p<0.001) but not men (p=0.069).

## Discussion

The present study sought to provide a comprehensive psychometric evaluation of the AASPIRE Social Support Scale, an autism-informed measure adapted through a CBPR process with autistic adults and other partners. Consistent with findings from the broader AASPIRE Measurement Toolkit validation study (Nicolaidis et al., 2026), the Social Support Scale demonstrated strong internal consistency overall and by reporter types (i.e., direct report , direct report with support, and caregiver report). Specifically, our findings support a multidimensional conceptualization of social support among autistic adults. The scale also demonstrated measurement invariance across reporter types. These findings suggest that the underlying construct of social support is measured similarly across reporters, supporting the use of the scale in heterogeneous samples that include autistic adults with varying support needs and communication abilities.

### Structural Validity

The five-factor structure provided strong empirical support for the multidimensional conceptualization that emerged from the AASPIRE measure-development process. The distinction among companionship, emotional support, informational support, and practical support is broadly consistent with the functional framework of social support proposed by Cohen and Wills (1985), while social isolation represents a related but distinct domain incorporated through the AASPIRE measure-development process.

The patterns of inter-factor correlations showed that the five factors were moderately to strongly correlated, suggesting that they reflect related but distinct aspects of social support. The particularly strong correlation between emotional and informational support may reflect overlap in the relationships through which these forms of support are experienced. Trusted and emotionally supportive relationships may also provide opportunities to seek information, advice, and guidance; alternatively, receiving useful information from another person may itself contribute to feeling understood and supported. In contrast, practical support demonstrated relatively modest correlations with the other support domains and the weakest association with social isolation. This finding suggests practical support may be less dependent on interpersonal connections than other forms of support. For instance, autistic adults may receive practical support from family members, caregivers, service providers, or formal support systems regardless of whether they experience a sense of belonging or social connection.

### Measurement Invariance

The AASPIRE Social Support Scale demonstrated measurement invariance across direct-report, direct-report with support, and caregiver-report groups, providing evidence that the underlying factor structure operates similarly across reporting methods. This is particularly important because different reporting approaches were intentionally designed to increase accessibility and accommodate autistic adults with diverse communication styles and support needs. Perceived social support is inherently subjective, making direct report from autistic adults important whenever possible; however caregiver report provides an alternative when direct reporting is not feasible. These findings support comparisons of social support across reporting approaches by reducing concern that observed group differences primarily reflect differences in how the underlying constructs are measured (Meredith, 1993). However, they should not be interpreted as demonstrating agreement between autistic adults and caregivers or equivalence of reporter perspectives. Future work could extend this finding by replication with larger caregiver samples and examination of demographic measurement invariance, which could further strengthen the evidence supporting the use of the AASPIRE Social Support Scale across diverse autistic populations.

### Group Differences in Social Support

After establishing measurement invariance, we observed substantial differences in social support across reporting types. Findings suggested that individuals completing the survey with support or through caregivers may have greater access to social, family, or service-related networks that facilitate broader support experiences. These differences may reflect characteristics of the autistic adults represented by each reporting approach. For instance, participants who required assistance with study participation may also have greater support needs in daily life and, in some cases, may be more likely to qualify for or receive formal services or assistance from caregivers. Importantly, greater support needs do not necessarily result in greater access to support, and autistic adults completing the survey independently may also experience unmet support needs or barriers to services. Future research should examine the extent to which observed scores differ across reporting methods and explore factors, such as daily living support needs and access to formal and informal support, that may contribute to reporter discrepancies.

The exploratory demographic comparisons also identified that younger autistic adults (18-29) experienced less support than older autistic adults. Given the limited research examining age-related differences in social support across the full adult lifespan in autism, the mechanisms underlying this finding remain unclear. Although transition-age literature has documented reduction in services and support following high school exit and elevated challenges establishing social relationships, employment, and community participation for young adults(Anderson et al., 2018; Roux et al., 2015), middle-age and older autistic adults continue to experience substantial challenges accessing support (Birtwell et al., 2026). These findings raise the possibility that social support networks, relationships, or access to particular forms of support may vary across adulthood.

Regarding gender differences, the finding that gender-diverse participants reported lower total social support than all men and women and greater isolation than women. Although these analyses were exploratory, the pattern is consistent with growing evidence that autistic transgender and gender-diverse individuals face unique social stressors and barriers to belonging (Kallitsounaki & Williams, 2023; Munday et al., 2025). No differences were identified by the broad BIPOC versus non-BIPOC categorization; however, this comparison combined heterogeneous racial and ethnic groups and should not be interpreted as evidence that race and ethnicity are unrelated to social support among autistic adults.

### Limitations and Future Directions

There are a few notable limitations of this study. First, while the large sample size allowed for a robust analytic approach involving CFA, a relatively small number of people used the caregiver report version of the scale (n=70). Replication in larger caregiver-report samples is therefore needed to strengthen the evidence supporting its use. Second, while the current study provides evidence that the underlying measurement structure is comparable across reporting approaches, we do not have paired self- and caregiver reports for the same autistic adults and therefore could not determine whether they produce similar scores for the same individual. Future studies using paired reports should examine agreement between autistic adults and caregivers. Third, reporting approach was associated with differences in participant characteristics, including characteristics likely to be related to support needs and access to support. Participants who completed the scale independently, with support, or through caregiver report represent populations that differ in communication characteristics, support needs, service use, and other factors related to social support. For example, the higher practical support in the caregiver-report group could reflect real differences in support needs and support availability rather than differences attributable to caregiver reporting. Future studies are needed to determine the extent to which observed differences are attributable to reporter perspective rather than differences in the populations represented by each reporting approach.

## Conclusion

The AASPIRE Social Support Scale provides a community-informed, multidimensional approach to assessing social support among autistic adults. Adapted from PROMIS measures through a CBPR process, the scale distinguishes the perceived availability of companionship, emotional, informational, and practical support while separately assessing subjective social isolation. Our findings support the factor structure, measurement invariance and internal consistency of the scale as a social support measure in autistic adult populations. Social support plays an important role in autistic adults’ mental well-being and overall quality of life (Charlton et al., 2023; Song et al., 2026) and is among the high-priority outcomes defined by autistic adults, researchers, and service providers (Nicolaidis et al., 2025). In addition to the rigorous testing, considerable efforts were made to ensure the measure’s understandability, ease of use, and relevance to autistic adults (Nicolaidis et al., 2026). As such, the AASPIRE Social Support Scale is an accessible, valid, and reliable tool that allows researchers, clinicians, providers, and policymakers to adequately assess the effectiveness of interventions.

## Supporting information

http://aaspire.org/wp-content/uploads/2026/04/AASPIRE-Social-Support-Scale.pdf

## Data Availability

All data from this study will be available through the National Institutes of Mental Health (NIMH) National Data Archive (NDA).

