## Supplementary material for "Development and Psychometric Evaluation of the AASPIRE Social Support Scale for Autistic Adults": http://aaspire.org/wp-content/uploads/2026/04/AASPIRE-Social-Support-Scale.pdf

Christina Nicolaidis, MD, MPH and the Outcomes Project Team

### AASPIRE Social Support Scale

*AASPIRE Social Support Scale Overview* 2
- Description 2
- Development 2
- Scoring 2
- Psychometric Properties 3
- Conditions for Use 3
- References 3

*AASPIRE Social Support Scale: Direct Report Version* 5
- Introduction 5
- Scale Items 6
  - Companionship 6
  - Emotional Support 7
  - Informational Support 8
  - Standard Direct Report items on perceived change and use of support to participate 12
- Hyperlinks 13

*AASPIRE Social Support Scale: Caregiver Report Version* 15
- Scale Items 16
  - Companionship 16
  - Informational Support 18
  - Practical Support 20
  - Social Isolation 22
  - Standard Caregiver Report items on perceived change and knowledge sources 23

*Detailed Comparison Between Original and Adapted Version* 24

Copyright 2025, Academic Autism Spectrum
Partnership in Research and Education (AASPIRE)

### AASPIRE Social Support Scale Overview

#### Description

The AASPIRE Social Support Scale assesses companionship, emotional support, informational support, practical support, and isolation. It represents a major adaptation of the PROMIS Social Support scales (Hahn et al., 2014).

#### Development

The AASPIRE Outcomes Project is using a community based participatory research (CBPR) approach to 1) create and validate the AASPIRE Measurement Toolkit; and 2) understand what predicts changes in outcomes over time (Nicolaidis et al., 2026a). Each instrument in the AASPIRE Measurement Toolkit has self- and caregiver-reported versions. AASPIRE developed and tested the instruments with and for autistic adults with or without intellectual disability, but they could also be used with general populations or other people who may need cognitively accessible survey instruments.
The AASPIRE Outcomes Project community-academic team collaboratively adapted this instrument for inclusion in the AASPIRE Measurement Toolkit.

Adaptations included adding prefaces to each subscale; adding visuals to all response options (cylinders filled to different proportions); modifying text to clarify terms and concepts; and substituting some items with other items from the PROMIS item banks or with new items we created to better capture autism-related dimensions of social well-being. Additionally, for the Practical Support subscale, we added hotlinks with more detailed instructions for how to answer if participants do or do not currently need support for the activities described in the items; and we added a 5th item asking about whether someone would notice and step in if they needed help.
See below for a more detailed comparison between the original and adapted instrument.

#### Scoring

The Companionship, Emotional Support, Informational Support, and Practical Support sub-scales use a 5-point Likert scale with 1=Never; 2=Rarely; 3=Sometimes; 4=Usually; and 5=Always. The Social Isolation subscale uses a 5-point Likert scale with 5=Never; 4=Rarely; 3=Sometimes; 2=Usually; and 1=Always.

Copyright 2025, Academic Autism Spectrum Partnership in Research and Education (AASPIRE)

Subscales are scored as the sum of all items. The total Social Support Scale is the sum of the five sub-scales. In all cases, higher total scores indicate greater social support (or less isolation). Additional items are intended for clinical or research purposes and are not part of the total score.

#### Psychometric Properties

The AASPIRE Social Support Scale is being psychometrically tested as part of the AASPIRE Outcomes Study, a longitudinal cohort study following a heterogeneous sample of ~850 autistic adults over time (Nicolaidis et al., 2026a).

Cognitive interviews have demonstrated strong content validity. Survey data have demonstrated excellent internal consistency reliability (alpha=0.94) and 2-week test-retest reliability (intraclass coefficient of 0.91) for the total scale score and across Social Support sub-scales, including Companionship (alpha=0.89, intraclass coefficient of 0.89), Emotional Support (alpha=0.88, intraclass coefficient of 0.90), Informational Support (alpha=0.90, intraclass coefficient 0.81), Practical Support (alpha=0.92, intraclass coefficient of 0.91), and Isolation-subscales (alpha=0.87, intraclass coefficient of 0.89). All subscales also demonstrated strong convergent and discriminant validity and 6-month responsiveness to change (Nicolaidis et al., 2026b).

Additional results about the scale's structural and construct validity are pending publication and will be shared on the AASPIRE Measurement Toolkit site when available.

#### Conditions for Use

The AASPIRE Social Supports Scale is available for use with attribution in noncommercial, clinical, and research settings. Permission is required for instrument adaptation, modification, and for-profit use. For permissions inquiries, contact Christina Nicolaidis and AASPIRE at.

Nicolaidis, C., Moura, I., Baker-Ericzén, M., Scharer, M., Maslak, J., Kripke-Ludwig, R., Horner-Johnson, W., Love, J., Raymaker, D., Joyce, A., des Roches Rosa, S., Herbert, G., Lounds Taylor, J., Yang, L.-Q., McDonald, K., Siddeek, Z., Kapp, S. K., Frowner, E., Smith, I., Wallington, A., Flores, K. J.,

Copyright 2025, Academic Autism Spectrum Partnership in Research and Education (AASPIRE)

Koffman, N., Kripke, C., Vera, J., Grillo, V. D., & Gardiner, F. (2026). *Addressing common problems in autism research: Cohort development and sample description for a participatory, real-world, longitudinal study of outcomes in autistic adults.* PsyArXiv. https://osf.io/preprints/psyarxiv/4m3bf

Nicolaidis, C., Raymaker, D. M., Baker-Ericzén, M., des Roches Rosa, S., Edwards, T., Frowner, E., Horner-Johnson, W., Joyce, A., Kapp, S. K., Kripke, C., Lounds Taylor, J., Love, J., Kripke-Ludwig, R., Maslak, J., McDonald, K., Moura, I., Scharer, M., Siddeek, Z., Smith, Y., Vera, J., . . . Flores, K. J. (2026). *Measuring high-priority outcomes in autistic adults: Initial psychometric assessment of the instruments in the AASPIRE Measurement Toolkit.* MedRxiv. https://doi.org/10.64898/2026.03.23.26349108

Copyright 2025, Academic Autism Spectrum Partnership in Research and Education (AASPIRE)

### AASPIRE Social Support Scale: Direct Report Version

#### Introduction

The next 4 sets of questions ask about different types of support you may get from other people.

When answering these questions, remember:

- The support can come from **anyone in your life** – it can be from family members, friends, romantic partners, co-workers, caregivers, paid staff, therapists, or anyone else.
- It can come from one person or it can come from many people.
- Only think about the times when you **want or need** support.
- Only think about the support that you **like or find helpful**.
- We are **not** asking about how often you need support. Instead, we are asking if the support **would be there if you ever needed it**.

Please mark one answer per row.

Copyright 2025, Academic Autism Spectrum
Partnership in Research and Education (AASPIRE)

### Scale Items

#### Companionship

Only think about the times that you **want** to do any of the following things with another person. Imagine what these things mean for you. Think about the people you have in your life **now.**

| When you **want** it… | Never | Rarely | Sometimes | Usually | Always |
|---|---|---|---|---|---|
| How often do you have someone to **relax** with? | ○ | ○ | ○ | ○ | ○ |
| How often do you have someone to **do something enjoyable** with? | ○ | ○ | ○ | ○ | ○ |
| How often do you have someone to **spend time** with? | ○ | ○ | ○ | ○ | ○ |
| How often do you have someone to **celebrate special occasions** with? | ○ | ○ | ○ | ○ | ○ |

Overall, in the last six months, did your companionship...

- ○ Get worse
- ○ Stay about the same
- ○ Get better

Copyright 2025, Academic Autism Spectrum Partnership in Research and Education (AASPIRE)

#### Emotional Support

**Only** think about the times that you **want** any of the following things from another person. Think about the people you have in your life **now**.

| When you **want** it… | Never | Rarely | Sometimes | Usually | Always |
|---|---|---|---|---|---|
| How often do you have someone **who will listen to you** when you need to talk? | ○ | ○ | ○ | ○ | ○ |
| How often do you have someone **you can share personal information** with about yourself or your problems? | ○ | ○ | ○ | ○ | ○ |
| How often do you have someone who **makes you feel appreciated**? | ○ | ○ | ○ | ○ | ○ |
| How often do you feel there are people in your life who **really understand you**? | ○ | ○ | ○ | ○ | ○ |

Overall, in the last six months, did your emotional support...

- ○ Get worse
- ○ Stay about the same
- ○ Get better

Copyright 2025, Academic Autism Spectrum Partnership in Research and Education (AASPIRE)

#### Informational Support

When answering the following questions, only think about the times you **want** information, suggestions, or advice **from other people in your life**. Think about the people you have in your life **now**.

| When you **want** it… | Never | Rarely | Sometimes | Usually | Always |
|---|---|---|---|---|---|
| How often can you **get information you trust** from other people? | ○ | ○ | ○ | ○ | ○ |
| How often do you have **someone to turn to for suggestions** about how to deal with a problem? | ○ | ○ | ○ | ○ | ○ |
| When dealing with a problem, how often can you **get helpful advice** from other people? | ○ | ○ | ○ | ○ | ○ |
| How often can you get good advice from other people on **what to do about a crisis**? | ○ | ○ | ○ | ○ | ○ |

Overall, in the last six months, did your informational support...

- ○ Get worse
- ○ Stay about the same
- ○ Get better

Copyright 2025, Academic Autism Spectrum Partnership in Research and Education (AASPIRE)

#### Practical Support

The following set of questions ask how often you have someone to help you do things.
We are **NOT** asking how often you *need* help to do these things yourself.

No matter how often you need help, we are interested in how much support **you have - or think you *would* have - *if* you ever were to need it.**

Think about the people you have in your life now.

You can click these links to see examples of how different people may answer the questions if:

- They need support to do these things some of the time
- They don't need support to do these things now

| | Never | Rarely | Sometimes | Usually | Always |
|---|---|---|---|---|---|
| How often *would you* have someone to **make your meals** if you couldn't do it yourself? | ○ | ○ | ○ | ○ | ○ |
| How often *would you* have someone to **bring you to an appointment** if you couldn't go by yourself? | ○ | ○ | ○ | ○ | ○ |
| How often *would you* have someone to help you with your **daily chores** if you couldn't do them yourself? | ○ | ○ | ○ | ○ | ○ |

Copyright 2025, Academic Autism Spectrum Partnership in Research and Education (AASPIRE)

| | Never | Rarely | Sometimes | Usually | Always |
|---|---|---|---|---|---|
| How often *would you* have someone to run **errands** for you, if you couldn't do them yourself? | ○ | ○ | ○ | ○ | ○ |
| How often *would you* have someone to **notice and step in** if you needed help? | ○ | ○ | ○ | ○ | ○ |

Overall, in the last six months, did your practical support...

- ○ Get worse
- ○ Stay about the same
- ○ Get better

Copyright 2025, Academic Autism Spectrum Partnership in Research and Education (AASPIRE)

#### Social Isolation

The next set of questions asks about times where you may feel isolated from other people. Think about the people you have in your life **now.**

| | Never | Rarely | Sometimes | Usually | Always |
|---|---|---|---|---|---|
| How often do you feel **left out**? | ○ | ○ | ○ | ○ | ○ |
| How often do you feel that people **barely know you**? | ○ | ○ | ○ | ○ | ○ |
| How often do you feel **isolated** from others? | ○ | ○ | ○ | ○ | ○ |
| How often do you feel that you are **alone in your interests and ideas**? | ○ | ○ | ○ | ○ | ○ |

Overall, in the last six months, did your feelings of isolation...

- Get worse
- Stay about the same
- Get better

Copyright 2025, Academic Autism Spectrum Partnership in Research and Education (AASPIRE)

#### Standard Direct Report items on perceived change and use of support to participate

Did you get any help while answering the questions in this section of the survey?

- ○ Yes
- ○ No

*<If 'Yes' to get help question>*
What kind of help did you get while answering the questions in this section of the survey? (Check all that apply.)

- ❑ Someone helped me use a computer, smartphone, or other device (for example, they clicked on the answers I chose).
- ❑ Someone read the survey to me.
- ❑ Someone helped me understand what the questions or answers options meant.
- ❑ Someone helped me decide how to answer the questions on the survey.
- ❑ I got some other type of help to answer the questions.
- ❑ Someone answered the questions for me, without my input.

Copyright 2025, Academic Autism Spectrum Partnership in Research and Education (AASPIRE)

#### Hyperlinks

You can click these links to see examples of how different people may answer the questions if:

They need support to do these things some of the time.

Here are some examples of how different people may answer the questions about how much practical support they get:

- Bob normally needs someone to prepare his lunch and dinner. Bob doesn't need help with breakfast. He gets all the help he needs to make lunch and dinner, so he would answer "always."
- Akira often needs help preparing meals, but they rarely get the help they need. Akira would mark "rarely."
- Jose only needs help going to appointments a few times a year. When he needs help, he always gets it. Even though he rarely needs help, he would answer that he "always" gets it.

Copyright 2025, Academic Autism Spectrum Partnership in Research and Education (AASPIRE)

You can click these links to see examples of how different people may answer the questions if:

- They don't need support to do these things now.

Tanisha doesn't normally need any help to prepare her meals. But Tanisha could need help *if* she were really sick and had to stay in bed.

- Tanisha should **imagine** what **would** happen if she ever needed help preparing her meals.
- She thinks her sister would help her some of the time, but she worries that her sister wouldn't be able to help her all the time.
- She doesn't have anyone else in her life that would be able to help. She would answer "sometimes."
- She doesn't have to know for sure what would happen. **It's ok for her to just guess.**

Copyright 2025, Academic Autism Spectrum
Partnership in Research and Education (AASPIRE)

### AASPIRE Social Support Scale: Caregiver Report Version

#### Introduction

The next 4 sets of questions ask about different types of support they may get from other people.

When answering these questions remember:

- The support can come from **anyone in their life** – it can be from family members, friends, romantic partners, co-workers, caregivers, paid staff, therapists, or anyone else.
- It can come from one person or it can come from many people.
- Only think about the times when they **want or need** support.
- Only think about the support that they **like or find helpful**.
- We are **not** asking about how often you think they need support. Instead, we are asking if the support **would be there if they ever needed it**.

Please mark one answer per row.

Copyright 2025, Academic Autism Spectrum Partnership in Research and Education (AASPIRE)

#### Scale Items

##### Companionship

Only think about the times that you think **they want** to do any of the following things with another person. Imagine what these things mean for them. Think about the people they have in their life **now**.

| When you think they **want** it… | Never | Rarely | Sometimes | Usually | Always |
|---|---|---|---|---|---|
| How often do you think they have someone to **relax** with? | ○ | ○ | ○ | ○ | ○ |
| How often do you think they have someone to **do something enjoyable** with? | ○ | ○ | ○ | ○ | ○ |
| How often do you think they have someone to **spend time** with? | ○ | ○ | ○ | ○ | ○ |
| How often do you think they have someone to **celebrate special occasions** with? | ○ | ○ | ○ | ○ | ○ |

Overall, in the last six months, do you think their companionship...

- ○ Got worse
- ○ Stayed about the same
- ○ Got better

Copyright 2025, Academic Autism Spectrum Partnership in Research and Education (AASPIRE)

#### Emotional Support

**Only** think about the times that you think **they want** any of the following things from another person. Think about the people they have in their life **now**.

| When you think they **want** it... | Never | Rarely | Sometimes | Usually | Always |
|---|---|---|---|---|---|
| How often do you think they have someone **who will listen** to them when they need to talk (or communicate in other ways)? | ○ | ○ | ○ | ○ | ○ |
| How often do you think they have someone **they can share personal information** with about themselves or their problems? | ○ | ○ | ○ | ○ | ○ |
| How often do you think they have someone who **makes them feel appreciated**? | ○ | ○ | ○ | ○ | ○ |
| How often do you think they feel there are people in their life who **really understand them**? | ○ | ○ | ○ | ○ | ○ |

Overall, in the last six months, do you think their emotional support...

- Got worse
- Stayed about the same
- Got better

Copyright 2025, Academic Autism Spectrum Partnership in Research and Education (AASPIRE)

#### Informational Support

When answering the following questions, only think about the times you think they **want** information, suggestions, or advice **from other people in their life**. Think about the people they have in their life **now.**

| When you think they **want** it… | Never | Rarely | Sometimes | Usually | Always |
|---|---|---|---|---|---|
| How often do you think they can **get information they trust** from other people? | ○ | ○ | ○ | ○ | ○ |
| How often do you think they have **someone to turn to for suggestions** about how to deal with a problem? | ○ | ○ | ○ | ○ | ○ |
| When dealing with a problem, how often do you think they can **get helpful advice** from other people? | ○ | ○ | ○ | ○ | ○ |
| How often do you think they can **get good advice** from other people on what to do about a crisis? | ○ | ○ | ○ | ○ | ○ |

Overall, in the last six months, do you think their informational support...

- ○ Got worse
- ○ Stayed about the same
- ○ Got better

Copyright 2025, Academic Autism Spectrum Partnership in Research and Education (AASPIRE)

#### Practical Support

The following set of questions ask how often you think they have someone to help them do things. We are **NOT** asking how often you think they *need* help to do these things themselves.
No matter how often they need help, we are interested in how much support **you think they have - or think they *would* have - *if* they ever were to need it.**

Think about the people they have in their life **now.**

You can click here to see examples of how different people may answer the questions about practical support.

| | Never | Rarely | Sometimes | Usually | Always |
|---|---|---|---|---|---|
| How often do you think they would have someone to **make their meals** if they couldn't do it themselves? | ○ | ○ | ○ | ○ | ○ |
| How often do you think they would have someone to **bring them to an appointment** if they couldn't go by themselves? | ○ | ○ | ○ | ○ | ○ |
| How often do you think they would have someone to help them with their **daily chores** if they couldn't do them themselves? | ○ | ○ | ○ | ○ | ○ |

Copyright 2025, Academic Autism Spectrum Partnership in Research and Education (AASPIRE)

| | Never | Rarely | Sometimes | Usually | Always |
|---|---|---|---|---|---|
| How often do you think they would have someone to run **errands** for them, if they couldn't do them themselves? | ○ | ○ | ○ | ○ | ○ |
| How often do you think they would have someone to **notice and step in** if they needed help? | ○ | ○ | ○ | ○ | ○ |

Overall, in the last six months, do you think their practical support...

- Got worse
- Stayed about the same
- Got better

Copyright 2025, Academic Autism Spectrum Partnership in Research and Education (AASPIRE)

#### Social Isolation

The next set of questions asks about times where you think they may feel isolated from other people. Think about the people they have in their life **now.**

| | Never | Rarely | Sometimes | Usually | Always |
|---|---|---|---|---|---|
| How often do you think they feel **left out**? | ○ | ○ | ○ | ○ | ○ |
| How often do you think they feel that people **barely know them**? | ○ | ○ | ○ | ○ | ○ |
| How often do you think they feel **isolated** from others? | ○ | ○ | ○ | ○ | ○ |
| How often do you think they feel that they are **alone in their interests and ideas**? | ○ | ○ | ○ | ○ | ○ |

Overall, in the last six months, do you think their feelings of isolation...

- Got worse
- Stayed about the same
- Got better

Copyright 2025, Academic Autism Spectrum Partnership in Research and Education (AASPIRE)

#### Standard Caregiver Report items on perceived change and knowledge sources

Overall, in the last six months, did the things this survey asked about...

- ○ Get worse
- ○ Stay about the same
- ○ Get better

How do you know this information?

- ❑ They told me (in whatever mode they communicate)
- ❑ I have observed the action, behavior, or events
- ❑ I can interpret their behaviors and/or vocalizations
- ❑ Someone else told me
- ❑ I feel like I can guess because I have known them for a long time
- ❑ I feel like I can guess because I spend a lot of time with them
- ❑ Other *<write-in>*
- ❑ None of the above

On a scale of 0 to 10, how confident are you in your responses?
*<0 - Not at all Confident to 10 - Totally Confident>*

OPTIONAL: If you are having trouble answering these questions, you may use this space to further clarify or explain your answers. *<write-in>*

Copyright 2025, Academic Autism Spectrum Partnership in Research and Education (AASPIRE)

### Detailed Comparison Between Original and Adapted Version

Changes between the PROMIS Social Support Scale and Social Support Scale- AASPIRE

| | **Original** | **Revised** |
|---|---|---|
| **Introduction** | <No introduction included in original version> | The next 4 sets of questions ask about different types of support you may get from other people.<br>When answering these questions, remember:<br>• The support can come from **anyone in your life**–it can be from family members, friends, romantic partners, co-workers, caregivers, paid staff, therapists, or anyone else.<br>• It can come from one person or it can come from many people.<br>• Only think about the times when you **want or need** support.<br>• Only think about the support that you **like or find helpful**.<br>• We are **not** asking about how often you need support. Instead, we are asking if the support **would be there if you ever needed it**. |
| **Response Option #1** | Never | 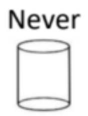  |

Copyright 2025, Academic Autism Spectrum Partnership in Research and Education (AASPIRE)

| | | |
|---|---|---|
| **Response Option #2** | Rarely | 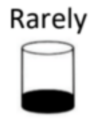 |
| **Response Option #3** | Sometimes | 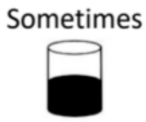 |
| **Response Option #4** | Usually | 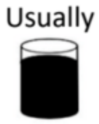 |
| **Response Option #5** | Always | 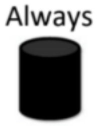 |

**Companionship**

| | **Original** | **Revised** |
|---|---|---|
| **Introduction** | <No introduction included in original version> | Only think about the times that you **want** to do any of the following things with another person. Imagine what these things mean for you. Think about the people you have in your life **now**. |
| **Item Change #1** | Do you have someone with whom to relax? | How often do you have someone to **relax** with? |
| **Item Change #2** | Do you have someone with whom you can do something enjoyable? | How often do you have someone to **do something enjoyable** with? |
| **Item Change #3** | Can you find companionship when you want it? | How often do you have someone to **spend time** with? |

Copyright 2025, Academic Autism Spectrum Partnership in Research and Education (AASPIRE)

| | | |
|---|---|---|
| **Removed Items** | Do you have someone with whom to have fun? | <No removed item in revised version> |
| **Added from Item bank** | Do you have someone with whom you can celebrate special occasions? | How often do you have someone **to celebrate special occasions** with? |

**Informational Support**

| | **Original** | **Revised** |
|---|---|---|
| **Introduction** | <No introduction in original version> | When answering the following questions, only think about the times you **want** information, suggestions, or **advice from other people in your life**. Think about the people you have in your life **now** |
| **Item Change #1** | I have someone to give me information if I need it. | How often can you **get information you trust** from other people? |
| **Item Change #2** | I have someone to turn to for suggestions about how to deal with a problem. | How often do you have **someone to turn to for suggestions** about how to deal with a problem? |
| **Item Change #3** | I get useful advice about important things in life. | When dealing with a problem, how often can you **get helpful advice** from other people? |
| **Item Change #4** | I have someone to give me good advice about a crisis if I need it. | How often can you get good advice from other people on **what to do about a crisis**? |

**Practical Support**

| | **Original** | **Revised** |
|---|---|---|
| **Introduction** | <No introduction included in original version> | The following set of questions ask how often you have someone to help you do things. We are **NOT** asking |

Copyright 2025, Academic Autism Spectrum Partnership in Research and Education (AASPIRE)

| | | |
|---|---|---|
| | | how often you *need* help to do these things yourself.<br><br>No matter how often you need help, we are interested in how much support **you have-or think you would have-*if* you ever were to need it**.<br><br>Think about the people you have in your life now. |
| **Additional Information** | <No additional information included in original version> | You can click these links to see examples of how different people may answer the questions if:<br><br>• They need support to do these things some of the time<br><br>• They don't need support to do these things now |
| **Item Change #1** | Do you have someone to take you to the doctor if you need it? | How often would you have someone **to bring you to an appointment** if you couldn't go by yourself? |
| **Item Change #2** | Do you have someone to help with your daily chores if you are sick? | How often would you have someone to help you with your **daily chores** if you couldn't do them yourself? |
| **Item Change #3** | Do you have someone to run errands if you need it? | How often would you have someone to run **errands** for you, if you couldn't do them yourself? |
| **Removed Item** | Do you have someone to help you if you are confined to bed? | <no removed item in revised version> |
| **Added from Item Bank** | Do you have someone to prepare your meals if you are unable to do it yourself? | How often would you have someone to **make your meals** If you couldn't do it yourself? |
| **Additional Item** | <No additional item in original version> | How often *would you* have someone **to notice and step in** if you needed help? |

Copyright 2025, Academic Autism Spectrum Partnership in Research and Education (AASPIRE)

#### Emotional Support

| | Original | Revised |
|---|---|---|
| **Introduction** | <No introduction in original version> | **Only** think about the times that you **want** any of the following things from another person. Think about the people you have in your life **now**. |
| **Item Change #1** | I have someone who will listen to me when I need to talk. | How often do you have someone **who will listen to you** when you need to talk? |
| **Item Change #2** | I have someone to confide in or talk to about myself or my problems. | How often do you have someone **you can share personal information** with about yourself or your problems? |
| **Item Change #3** | I have someone who makes me feel appreciated. | How often do you have someone who **makes you feel appreciated**? |
| **Removed Item** | I have someone to talk with when I have a bad day. | <No removed item in revised version> |
| **Added from Item Bank** | I feel there are people who really understand me | How often do you feel there are people in your life who **really understand you**? |

#### Social Isolation

| | Original | Adapted |
|---|---|---|
| **Introduction** | <No introduction in original version> | The next set of questions asks about times where you may feel isolated from other people. Think about the people you have in your life **now**. |
| **Item Change #1** | I feel left out. | How often do you feel **left out**? |

| | Original | Adapted |
|---|---|---|
| **Item Change #2** | I feel that people barely know me. | How often do you feel that people **barely know you**? |
| **Item Change #3** | I feel Isolated from others. | How often do you feel **Isolated** from others? |
| **Removed Item** | I feel that people are around me but not with me. | <No removed item in revised version> |
| **Added from Item Bank** | I feel that I am alone in my interests and ideas | How often do you feel that you are **alone in your interests and ideas**? |

Copyright 2025, Academic Autism Spectrum Partnership in Research and Education (AASPIRE)
